# A Scoping Review of Published and Ongoing Prospective Meta-Analyses in Health Research: Study Protocol

**DOI:** 10.1101/2023.05.18.23290213

**Authors:** Thomas Love, Xiang Li, James X. Sotiropoulos, Jonathan G. Williams, Sol Libesman, Kylie E. Hunter, Anna Lene Seidler

## Abstract

**Background:** Prospective meta-analysis (PMA) is an emerging evidence synthesis methodology, in which the study selection criteria, hypotheses and analyses are finalised prior to the knowledge of included study results. This has numerous benefits over retrospective meta-analysis, including a reduction in selection bias, selective outcome reporting and publication bias and improved data harmonisation. Yet, common misunderstandings of the PMA methodology remain, leading to inappropriate and/or suboptimal implementation by researchers.

**Objectives:** To investigate the landscape of the published and ongoing prospective meta-analyses.

**Methods:** We will systematically search the International prospective Register of Systematic Reviews (PROSPERO) for PMAs from inception to January 2023 and screen these studies in COVIDENCE. The results of this search will be combined with a search of medical databases (PubMed, Embase, Cochrane Database of Systematic Reviews) up to 2018, to capture PMA that started prior to the launch and subsequently increasingly wide implementation of PROSPERO in February 2011. All PMAs conducted in human health research will be eligible for inclusion. All records will be double-screened, and data will be double extracted, with conflicts resolved through consensus.

**Dissemination:** Insights from this scoping review will inform adaptions to current PMA guidelines and thus assist researchers in planning future PMAs in their fields.

## Introduction

Systematic reviews and meta-analyses involve the synthesis of data from multiple sources to increase statistical power and improve the generalisability of results [1]. Most meta-analyses are conducted retrospectively, where the planning of analysis, selection of studies and aggregation of data generally occurs following the publication of individual study results [2]. This prior knowledge of results of included studies may introduce bias into both study selection and meta-analysis methods, potentially influencing the meta-analysis hypotheses, inclusion criteria and outcomes [2]. In addition, retrospective meta-analyses are prone to publication bias, whereby studies with positive or statistically significant results are more likely to be published and thus included in meta syntheses [2,3]. Reviewer selection bias is also prevalent, since reviewers know the results of existing studies and may shape their selection criteria (inadvertently or deliberately) to include studies that confirm their pre-existing beliefs and exclude those that do not [4]. Further, selective outcome reporting frequently occurs in human health research [5-7], involving only a subset of the original recorded variables being selected for inclusion in the final publication based on knowledge of the results. This may lead to bias in subsequent meta-analyses that include these studies, reducing trustworthiness in their findings [6, 8].

Prospective meta-analysis (PMA) is a “next-generation” methodology that addresses many of these limitations of retrospective meta-analysis [9]. PMA is defined as a meta-analysis in which the study selection criteria, hypotheses and analyses are finalised prior to the knowledge of included studies’ results [10-12]. Due to the prospective selection of studies, PMA can reduce the risks of selective outcome reporting and publication bias [13]. Further, specification of study criteria and outcomes prior to knowledge of results allows a reduction in selection bias [14]. PMAs are particularly advantageous to inform decision making for high priority research questions for which limited previous evidence exists and new data are expected to emerge rapidly, for example, during the COVID-19 pandemic [15, 16]. PMAs are usually highly collaborative and are ideally conducted by a central steering and data analysis committee, as well as representatives from each individual study [10].

PMAs allow harmonisation of core collected variables, including core outcomes time points and measures between each study, enabling successful synthesis of each study [14]. Harmonisation can increase the statistical power for detecting intervention effects, which is especially beneficial for rare but important outcomes such as mortality [13, 17]. For example, in a meta-analysis case study of early childhood obesity prevention, the proportion of outcome categories that could be meta-analysed due to inclusion in all eligible studies increased from 18% to 91% post-PMA inclusion [13]. Outcome harmonisation can thus reduce research waste [14, 17, 18]. However, some variation across included studies in context (protocol, populations, and interventions) is beneficial to increase the external validity of results [10].

PMAs can be either conducted with aggregate or individual participant data (IPD), albeit IPD PMAs are common due to the pre-existing collaborative structure facilitating collection of IPD [19]. Collecting IPD has the important advantage of facilitating subgroup analyses that are less prone to ecological bias [14]. In an IPD PMA this is further enhanced by the ability to prospectively harmonise a collection of subgroup variables across all studies [14].

PMAs can be planned at any point prior to results being known for the included studies. Thus, the possible timepoints of study inclusion are: during the early planning stages of individual studies, where recruiting/intervention delivery have already initiated, or even following the data collection of each study, however, prior to knowledge of results. Generally, the earlier a PMA is started the greater the ability to harmonise protocols, including core outcomes and subgroups [2, 10].

In some cases, PMA methodology can be combined with retrospective meta-analysis methodology, and this is called a nested PMA [10]. Prospective studies can be harmonised with each other and with the included retrospective studies to optimise data availability and synthesis. The comparison between prospectively included evidence and retrospective evidence in a sensitivity analysis allows for the assessment of publication bias and selective reporting bias [10].

**Figure 1.**
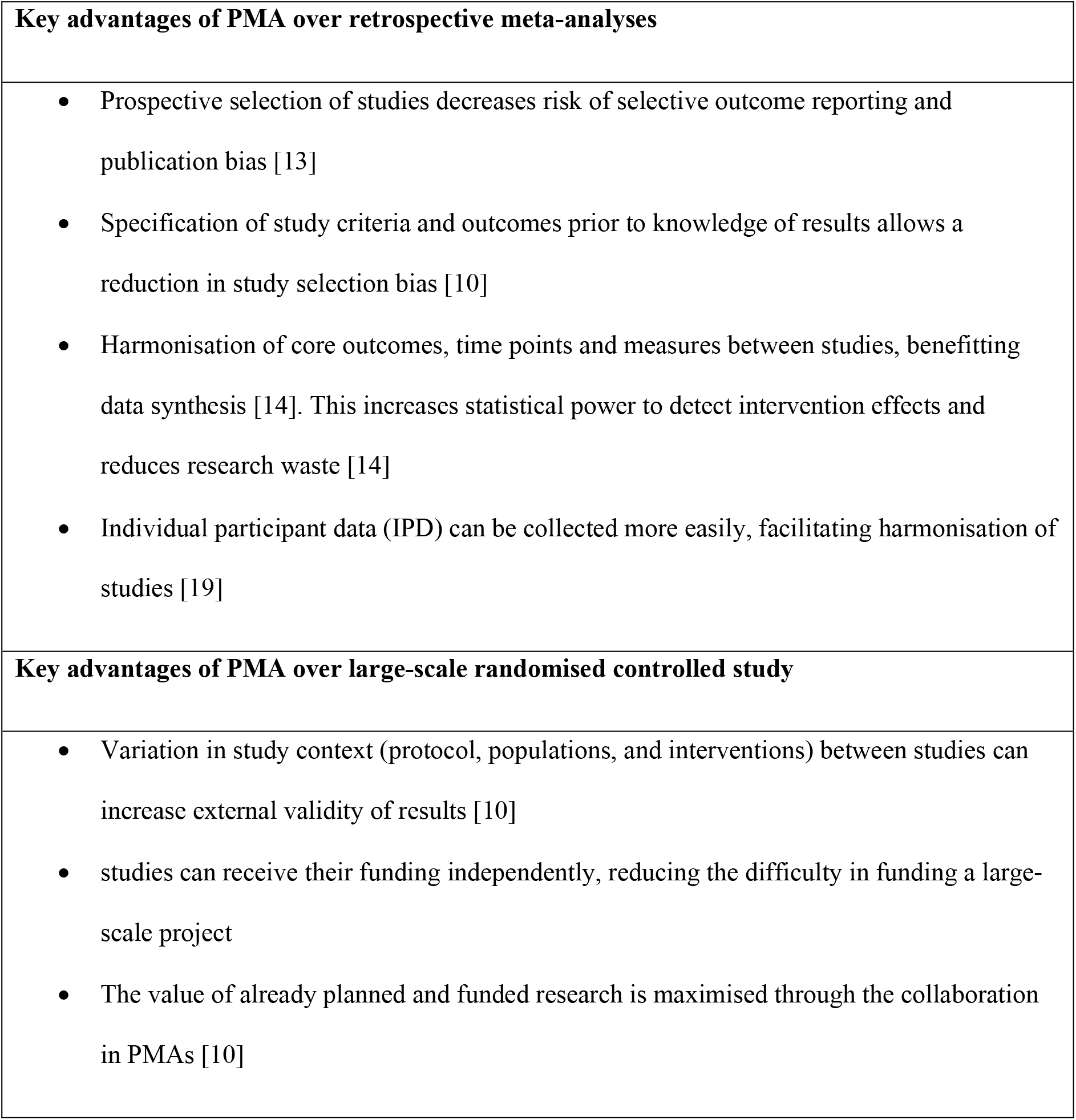
Key advantages of PMA over other research methods

Yet, there remain common misunderstandings of the methodology, with inappropriate and/or suboptimal implementation by researchers. Therefore, continued refinement of methods guidelines is required to optimise its future implementation and impact on scientific research. To progress this development, it is important to gain an overview of the current prevalence and type of PMAs being conducted. These insights will inform adaptions to the current PMA guidelines and assist researchers in planning PMAs in their fields.

We aim to:

- Describe and characterise the landscape of PMAs (ongoing or completed) within human health research
- Identify the questions that are being addressed by PMA methodology in human health research (including study design, populations and interventions)
- Describe the characteristics of PMAs (e.g. types, levels and forms of collaboration, number of participants and analysis types) that have been or are currently being conducted

## Methods

This protocol is reported in accordance with PRISMA-P (Preferred Reporting Items for Systematic review and Meta-Analysis Protocols) guidelines (appendix 1) and results will be reported in accordance with PRISMA-scoping review extension [20].

### Eligibility criteria

We are planning to include all PMAs that are conducted in human health research and fulfill PMA criteria as described in the literature [10]:

- Utilise any study design (interventional and observational studies)
- Are conducted in humans (excluding animal studies)
- Either use IPD or aggregate data
- Study an outcome associated with human health

#### Box 1.

Definition of a prospective meta-analysis [10] The key feature of a prospective meta-analysis (PMA) is that the studies or cohorts are identified as eligible for inclusion in the meta-analysis, and hypotheses and analyses are specified, before the results of the studies or cohorts related to the PMA research question are known.

### Information sources

An extensive literature search of ongoing and completed records on the International prospective Register of Systematic Reviews (PROSPERO) [21] will be conducted to identify PMAs. Additionally, Pubmed, Embase and the Cochrane Database of Systematic Reviews will also be searched.

### Search strategy

Firstly, we will search PROSPERO for any ongoing or completed PMAs registered from the establishment of the database (February 2011) to 1^st^ January 2023. All studies registered between these dates for which investigators ticked the box ‘prospective meta-analysis’ were exported to Covidence [22]. Further, we will conduct the following free text searches:

- Prospective meta-analysis
- Collaborat* meta-analysis
- Preplanned and meta-analysis
- Pre-planned and meta-analysis
- Prospectively planned meta-analysis

Secondly, the results of this search will be combined with a more comprehensive literature search conducted in February 2018 [23]. This included a search of PubMed (1950 to February 2018), Embase (1974 to February 2018), Cochrane Database of Systematic Reviews (all entries up to February 2018) and PROSPERO (inception to February 2018). Additionally, web searches were performed to identify collaborative PMA that may not have been published in traditional journals, and experts in the field (such as the Cochrane PMA methods group) were contacted to name additional planned or conducted PMAs known to them. The search terms and results of the 2018 search are attached in Appendix 2. We expect that most PMAs occurring after the search in February 2018 will appear in the updated PROSPERO search, due to the widespread adoption of PROSPERO registration for evidence synthesis projects. In addition, the references of identified relevant publications will be searched.

### Data management and selection process

The identified studies from the updated search will be exported as RIS files and the duplicates will be removed. A pilot screen will be conducted, and the review team will develop and test screening questions using Covidence. The selected PMA registrations will be screened by two authors (TL and XL). Both TL and XL will screen the records independently, with any differences being resolved by consensus and in consultation with JS and/or ALS. Identified studies that contain PMA keywords, however, do not satisfy the essential PMA criteria will be recorded. As the next steps, we will attempt to retrieve publications for all included PMA protocols. SCOPUS will be searched for these publications, beginning with PROSPERO registration number, and then moving on to study title and authors. Where available, we will then screen these full texts in duplicate, resolving conflicts by consensus and consultation.

### Data collection process, data items and categorisation of studies

Information on the key PMA elements and characteristics will be recorded in a structured table for each study, as shown in Table 1. This information includes relevant study data (e.g. study design, interventions and eligible populations) and also PMA specific details (e.g. type of PMA, level and structure of collaboration and publication policies). Extractions will occur in duplicate.

**Table 1.** Data to be extracted and collated from identified PMAs

| **Category** | **Data to be extracted** |
| --- | --- |
| General study information | Title |
|  | Authors |
|  | Registration year on PROSPERO |
|  | Results publication year (if applicable) |
|  | Follow-up results publication year (if applicable) |
|  | PROSPERO registration number |
|  | Most recent publication type |
|  | Journal |
|  | Studied Health Condition (WHO International Classification of Diseases) |
|  | Primary Purpose (Treatment: drugs, other, Prevention, Diagnosis) |
|  | Number of included studies |
|  | Number of participants |
|  | Is the PMA ongoing or are results available? |
| Review information | Eligible study designs |
|  | Eligible participant populations |
|  | Eligible interventions and comparators? |
|  | Outcome information |
|  | Region of PMA collection |
|  | Region of included studies in PMA |
|  | Funding |
| PMA-specific information | Type of PMA |
|  | Nested PMA |
|  | If systematic review was conducted prior to PMA |
|  | Included PRISMA checklist |
|  | How were individual studies identified? (What search method was used to identify relevant studies?) |
|  | Individual participant data? |
|  | Subgroup analyses performed? |
|  | Harmonization of outcomes |
|  | Quality of the primary study is assessed by which tools? |
| Collaboration management | Was there a collaboration? |
|  | Project management structures? |
|  | Procedures for data management? |
|  | Nominated statistical analysis group? |
| Publication policy | Authorship policy? |
|  | Writing committee? |
|  | Manuscript policy? |

The extraction records of the previous 2018 search [23] will be checked by a single reviewer. If any information in Table 1 is not available in the historical extraction record, two reviewers will update the extraction.

### Data synthesis

The total number of eligible studies from inception to 1^st^ January 2023 that fulfil PMA criteria will be recorded. The PMA details will be analysed both quantitatively (e.g. median sample size, numbers per PMA type), and qualitatively (e.g. describing the varying collaboration structures and common themes across collaborations/interesting case studies). The analysed study characteristics of confirmed and uncertain PMAs will be compared.

## Data Availability

N/A - protocol only

## Acknowledgements

Lisa Askie

Saskia Cheyne

## Author contributions

Thomas Love and Xiang Li are the co-first authors, contributing most of the work and they will conduct the review. James X. Sotiropoulos, Jonathan G. Williams, Sol Libesman and Kylie E. Hunter provided assistance and guidance in planning this review. Dr Anna Lene Seidler is the project supervisor, overseeing the project.

## Amendments to this protocol

This protocol will be uploaded to Open Science Framework. Important protocol amendments will be updated in Open Science Framework and recorded in the final report.

## Appendix 1 PRISMA-P checklist

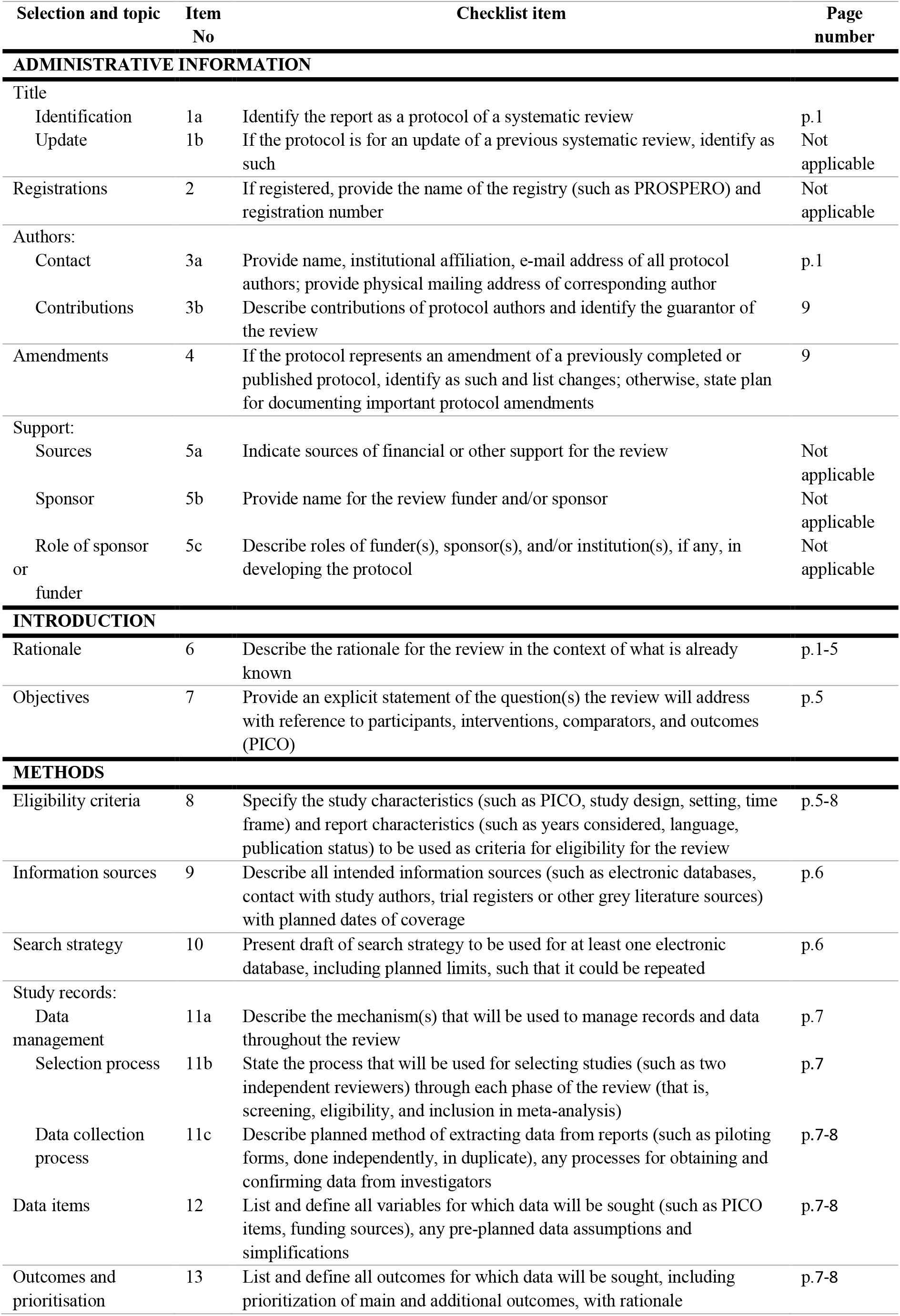
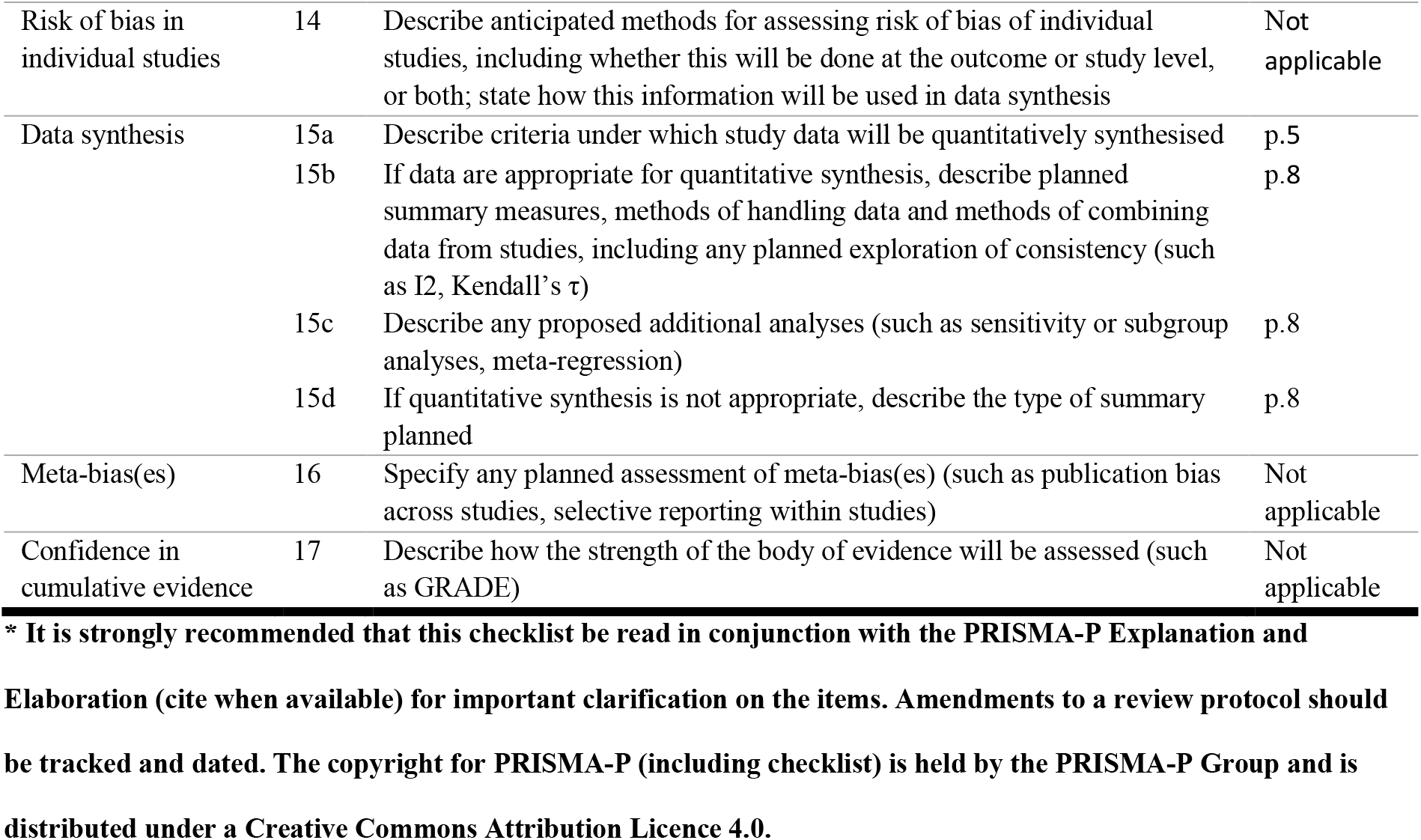

From: Shamseer L, Moher D, Clarke M, Ghersi D, Liberati A, Petticrew M, Shekelle P, Stewart L, PRISMA-P Group. Preferred reporting items for systematic review and meta-analysis protocols (PRISMA-P) 2015: elaboration and explanation. BMJ. 2015 Jan 2;349(jan02 1):g7647.

## Appendix 2 Previous search strategy

An extensive literature search was conducted to identify PMAs and PMA methodological reports. An incomplete list of known PMAs on the Cochrane PMA Methods Group website was carefully examined (http://methods.cochrane.org/pma/list-prospective-meta-analysis-studies) to identify any terminology used to describe PMAs. In addition, the researchers used their experience as convenors and editors of the Cochrane PMA Methods Group and authors of multiple previous PMAs to derive further search terms. The derived search strategy involved various combinations of the following terms, searching in abstracts and titles: prospective, combined, planned, preplanned, pre-planned, pooled, pre-specified, prespecified, collabo*, individual participant data, individual patient data, meta-analysis, meta analysis, metaanalysis, consortium.

### Previous search results

**Figure 1.**
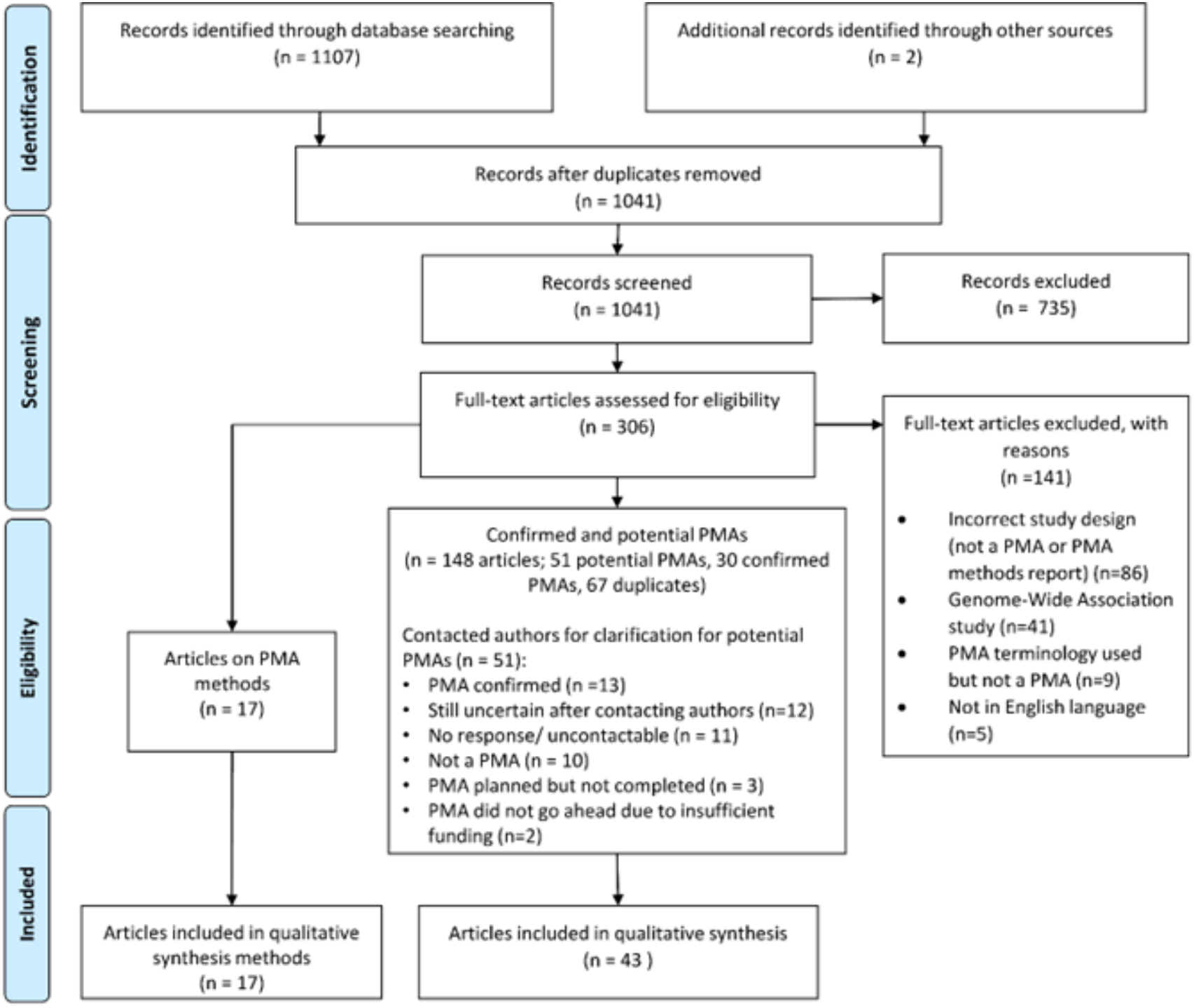
PRISMA flowchart

## References

1. Ahn, E. and H. Kang, Introduction to systematic review and meta-analysis. Korean J Anesthesiol, 2018. 71(2): p. 103–112.

2. Tierney, J.F., et al., A framework for prospective, adaptive meta-analysis (FAME) of aggregate data from randomised trials. PLoS Med, 2021. 18(5): p. e1003629.

3. Roberts, I. and K. Ker, How systematic reviews cause research waste. Lancet, 2015. 386(10003): p. 1536.

4. Ahmed, I., A.J. Sutton, and R.D. Riley, Assessment of publication bias, selection bias, and unavailable data in meta-analyses using individual participant data: a database survey. BMJ, 2012. 344: p. d7762.

5. Vassar, M., et al., Evaluation of selective outcome reporting and trial registration practices among addiction clinical trials. Addiction, 2020. 115(6): p. 1172–1179.

6. Riemer, M., et al., Trial registration and selective outcome reporting in 585 clinical trials investigating drugs for prevention of postoperative nausea and vomiting. BMC Anesthesiology, 2021. 21(1): p. 249.

7. Mathieu, S., et al., Comparison of registered and published primary outcomes in randomized controlled trials. Jama, 2009. 302(9): p. 977–84.

8. Page, M.J., et al., Bias due to selective inclusion and reporting of outcomes and analyses in systematic reviews of randomised trials of healthcare interventions. Cochrane Database Syst Rev, 2014. 2014(10): p. Mr000035.

9. Ioannidis, J., Next-generation systematic reviews: prospective meta-analysis, individual-level data, networks and umbrella reviews. British Journal of Sports Medicine, 2017. 51(20): p. 1456–1458.

10. Seidler, A.L., et al., A guide to prospective meta-analysis. BMJ, 2019. 367: p. 5342.

11. Reade, M.C., et al., Prospective meta-analysis using individual patient data in intensive care medicine. Intensive Care Medicine, 2010. 36(1): p. 11–21.

12. Chalmers, I., The Cochrane collaboration: preparing, maintaining, and disseminating systematic reviews of the effects of health care. Ann N Y Acad Sci, 1993. 703: p. 156–63; discussion 163-5.

13. Seidler, A.L., et al., Quantifying the advantages of conducting a prospective meta-analysis (PMA): a case study of early childhood obesity prevention. Trials, 2021. 22(1): p. 78.

14. Seidler, A.L., et al., Prospective meta-analyses and Cochrane’s role in embracing next-generation methodologies. Cochrane Database Syst Rev, 2020. 10(10): p. Ed000145.

15. Naci, H., et al., Producing and using timely comparative evidence on drugs: lessons from clinical trials for covid-19. BMJ, 2020. 371: p. m3869.

16. Sterne, J.A.C., et al., Association Between Administration of Systemic Corticosteroids and Mortality Among Critically Ill Patients With COVID-19: A Meta-analysis. Jama, 2020. 324(13): p. 1330–1341.

17. Efthimiou, O., Practical guide to the meta-analysis of rare events. Evid Based Ment Health, 2018. 21(2): p. 72–76.

18. Jonathan J Deeks, J.P.H., Douglas G Altman; on behalf of the Cochrane Statistical Methods Group. Chapter 10: Analysing data and undertaking meta-analyses Cochrane Handbook for Systematic Reviews of Interventions version 6.3 2019 February 2022; Available from: https://training.cochrane.org/handbook/current/chapter-10.

19. Reade, M.C., et al., Prospective meta-analysis using individual patient data in intensive care medicine. Intensive Care Med, 2010. 36(1): p. 11–21.

20. PRISMA Extension for Scoping Reviews (PRISMA-ScR): Checklist and Explanation Annals of Internal Medicine, 2018. 169(7): p. 467–473.

21. (NIHR), N.I.f.H.R., PROSPERO: International prospective register of systematic reviews. University of York: York, UK.

22. Cochrane. Covidence. Available from: https://www.covidence.org/.

23. Seidler, A.L., et al., A scoping review of prospective meta-analyses in health research T.U.o. Sydney, Editor. 2018.

